# Clinical and dosimetric dataset of time-to-event normal tissue complication probability for osteoradionecrosis

**DOI:** 10.1101/2025.08.12.25333524

**Authors:** Natalie A. West, Serageldin Kamel, Andrew Wentzel, Zaphanlene Kaffey, Moamen Abdelaal, G. Elisabeta Marai, Guadalupe Canahuate, Xinhua Zhang, Melissa M. Chen, Kareem A. Wahid, Jillian Rigert, Kristy K. Brock, Mark Chambers, Adegbenga O. Otun, Ruth Aponte-Wesson, Renjie He, Mohamed A. Naser, Katherine A. Hutcheson, Abdallah S. R. Mohamed, Lisanne V. van Dijk, Amy C. Moreno, Stephen Y. Lai, Clifton D. Fuller, Laia Humbert-Vidan

**Author notes:** **Corresponding author emails:** Natalie A. West, Clifton D. Fuller, Laia Humbert-Vidan. Spatial-Non-spatial Multi-Dimensional Analysis of Radiotherapy Treatment/Toxicity Team (SMART3) (Andrew Wentzel, G. Elisabeta Marai, Guadalupe Canahuate, Xinhua Zhang, Abdallah S. R. Mohamed, Clifton D. Fuller). **Public access policy compliance** In accordance with NOT-OD-25-049, Supplemental Guidance to the 2024 NIH Public Access Policy: Government Use License and Rights: “This manuscript is the result of funding in whole or in part by the National Institutes of Health (NIH). It is subject to the NIH Public Access Policy. Through acceptance of this federal funding, NIH has been given a right to make this manuscript publicly available in PubMed Central upon the Official Date of Publication, as defined by NIH.”. **Pre-print Statement** Consistent with NOT-OD-17-050, Reporting Preprints and Other Interim Research Products, as “NIH encourages investigators to use interim research products, such as preprints, to speed the dissemination and enhance the rigor of their work”, a pre-peer reviewed deposition of the initial submission version of the manuscript has been deposited for public access at medRxiv with DOI available upon acceptance. **Reporting Guideline Compliance Statement** In accordance with EQUATOR Network (Enhancing the QUAlity and Transparency Of health Research) guidance, we have utilized the RECORD Statement extended from the STROBE Statement; the completed checklist is attached as Supplementary file.

## Abstract

Osteoradionecrosis of the jaw (ORNJ) is a radiation-induced late toxicity that can dramatically decrease patients’ quality of life. Recent increases in survival rates of head and neck cancers associated with human papillomavirus (HPV) infection have resulted in a higher frequency of radiation-induced toxicities, particularly ORNJ. Recent work with Normal Tissue Complication Probability (NTCP) models and a Weibull Accelerated Failure Time (WAFT) model have further developed our understanding of ORNJ clinical/dosimetric risk factors and longitudinal features, respectively. In this data descriptor, 1129 head and neck cancer (HNC) patients received curative intent radiotherapy (RT) at MD Anderson Cancer Center and were follow-up with clinical and radiological assessments at 3-6, 12, 18, 24 months, and then annually following the conclusion of RT for development of ORNJ. This data, in addition to the patients’ demographic, supplementary clinical, and dosimetric information was recorded in a comma-separated value file embedded within this data descriptor. This large, longitudinal dataset is a significant resource for further systematic analysis of post-RT normal tissue outcomes in HNC.

## Background and Summary

Head and neck cancers (HNC) affect over 58,000 Americans annually, with a growing proportion attributed to human papillomavirus (HPV)^1^ infection. HPV-associated HNC are notably diagnosed in younger populations and are associated with higher survival rates in comparison to HPV-negative HNC^2^. Radiation therapy (RT) remains the mainstay of treatment for HPV-positive HNC, but the combination of RT with extended survival has led to an increased incidence of RT-induced late toxicities in normal tissues. One such complication is osteoradionecrosis of the jaw (ORNJ), a severe sequela following RT with an incidence ranging from 4 to 15%^3^. The mechanism of ORNJ is believed to be first instigated by compromised vascularity through hypoxic, hypovascular, and hypocellular tissue (Marx’s 3 H’s)^4^ followed by progressive loss in cortical bone integrity, ultimately impairing oral function and quality of life^5,6^. Due to the favorable RT response and prognosis of HPV-associated HNC and the subsequent number of patients transitioning to survivorship, there is a need to better understand the timing and progressive risk of ORNJ in relation to radiation treatment of HNC in order to optimize prevention efforts of this often debilitating condition.

Previous cross sectional statistical analyses^7–10^, including Normal Tissue Complication Probability (NTCP) models of ORNJ^11,12^, have identified clinical and dosimetric risk factors associated with this sequela. Additionally, some studies have explored statistical correlations on longitudinal ORNJ data^13,14^. In recent work, we developed a fully parametric multivariable Weibull Accelerated Failure Time (WAFT) model to predict patient-specific ORNJ risk over time based on longitudinal data.

This data descriptor presents the underlying dataset used for the development of the ORNJ WAFT model^15^. The dataset is comprised of a large, longitudinal cohort of HNC patients and includes detailed demographic, clinical, and dosimetric variables, along with structured follow-up data and time-to-ORNJ events. The availability of this dataset offers a valuable resource for modeling ORNJ and supports the development of predictive tools for personalized survivorship care in HNC.

## Methods

### IRB Protocol

Data for this retrospective acquisition received institutional review board (IRB) approval (RCR030800) and was collected from a philanthropically funded observational cohort (Stiefel Oropharynx Cancer Cohort, PA14-0947). All patients included were consented RT cases.

### Patient Population

1129 HNC patients from an internal MD Anderson Cancer Center cohort were treated with curative intent RT from 2005 to 2022. Patients were closely followed via clinical and radiological assessments every 3, 6, 12, 18, and 24 months, and then approximately annually following the conclusion of RT. Patient data was stored in and accessed via the Epic Electronic Health Record System.

### Demographic Data

All demographic, clinical, and dosimetric variables are summarized in **Table 1**. The patients’ demographic data included: gender (male or female), age (in years), smoking status (current, former, never), and smoking pack-years. Smoking pack-years were calculated by the product of tobacco packs smoked per day and number of years smoked.

**Table 1.**
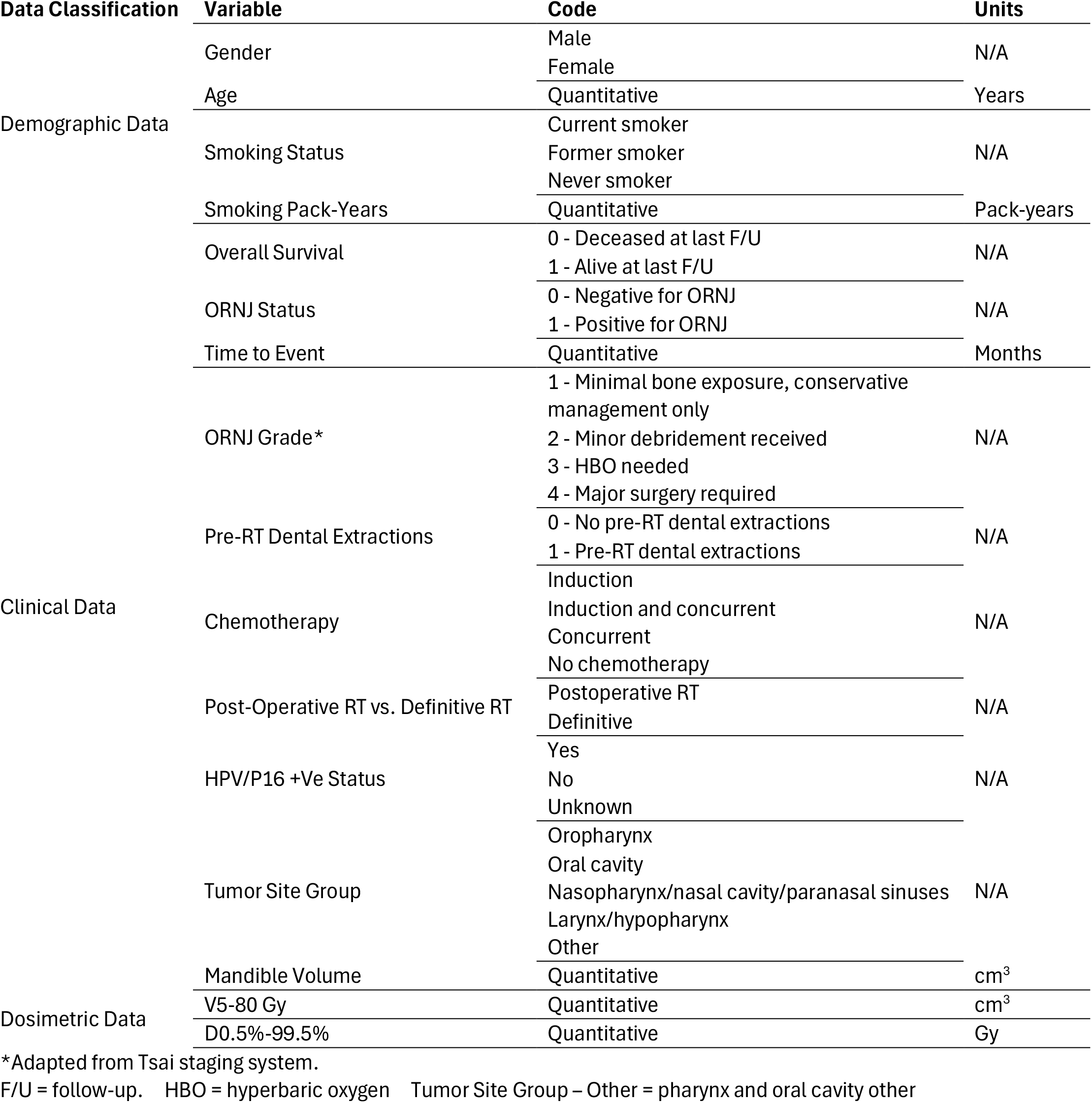
Demographic, clinical, and dosimetric data for included patient cohort showing variable, its respective coding (qualitative or quantitative) and units (when applicable). This can be used to supplement the CSV file included within this data descriptor.

**Table 2.** Distribution of demographic and clinical data stratified by ORNJ status (control group and ORNJ group). Columns three and four demonstrate n number of patients represented per variable in each population (control or ORNJ) and their relative percent distribution.

| Variable |  | Amount of Control (n=931) | Amount of ORNJ (n=198) |
| --- | --- | --- | --- |
| Gender | Male | 764 (82.1%) | 174 (87.9%) |
|  | Female | 167 (17.9%) | 24 (12.1%) |
| Median Age (IQR) |  | 60.0 (14.0) | 60.0 (12.0) |
| Primary Tumor Site | Oropharynx | 602 (64.7%) | 138 (69.7%) |
|  | Oral Cavity | 156 (16.8%) | 49 (24.8%) |
|  | Larynx/Hypopharynx | 187 (20.1%) | 7 (3.5%) |
|  | Nasopharynx, Nasal Cavity, Paranasal Sinuses | 21 (2.3%) | 2 (1.0%) |
|  | Other | 5 (0.5%) | 2 (1.0%) |
| T Stage | T0 | 33 (3.5%) | 3 (1.5%) |
|  | T1 | 210 (22.6%) | 31 (15.7%) |
|  | T2 | 314 (33.7%) | 63 (31.8%) |
|  | T3 | 200 (21.5%) | 39 (19.7%) |
|  | T4 | 160 (17.2%) | 59 (29.8%) |
|  | T4a | 8 (0.9%) | 3 (1.5%) |
|  | T4b | 0 (0.0%) | 0 (0.0%) |
|  | TX | 6 (0.6%) | 0 (0.0%) |
| N Stage | N0 | 189 (20.3%) | 32 (16.2%) |
|  | N1 | 210 (22.6%) | 30 (15.2%) |
|  | N2 | 205 (22.0%) | 69 (34.9%) |
|  | N2a | 23 (2.5%) | 2 (1.0%) |
|  | N2b | 181 (19.4%) | 40 (20.2%) |
|  | N2c | 99 (10.6%) | 21 (10.6%) |
|  | N3 | 21 (2.3%) | 4 (2.0%) |
|  | NX | 3 (0.3%) | 0 (0.0%) |
| HPV+ | Yes | 174 (18.7%) | 20 (10.1%) |
|  | No | 17 (1.8%) | 2 (1.0%) |
|  | Unknown | 740 (79.5%) | 176 (88.9%) |
| Smoking Status | Current | 126 (13.5%) | 33 (16.7%) |
|  | Former | 445 (47.8%) | 93 (47.0%) |
|  | Never | 360 (38.7%) | 72 (36.4%) |
| Pre-RT Dental Extraction* |  | 231 (24.8%) | 76 (38.4%) |
| Pre-RT Surgery | Postop RT | 154 (16.5) | 43 (21.7%) |
|  | Definitive RT | 777 (83.5%) | 155 (78.3%) |
| RT Technique | IMRT | 678 (72.8%) | 161 (81.3%) |
|  | VMAT | 188 (20.2%) | 24 (12.1%) |
|  | IMPT | 17 (1.8%) | 4 (2.0%) |
|  | Non-IMRT | 11 (1.2%) | 0 (0.0%) |
|  | N/A | 37 (4.0%) | 9 (4.6%) |
| Chemotherapy | Concurrent chemotherapy | 465 (50.0%) | 97 (49.0%) |
|  | Induction and concurrent | 202 (21.7%) | 59 (29.8%) |
|  | No chemotherapy | 189 (20.3%) | 24 (12.1%) |
|  | Induction chemotherapy | 75 (8.1%) | 18 (9.1%) |
| Median follow up time in years (range) |  | 8.0 (6.6) | 8.0 (5.4) |

\*Only dental extractions within six weeks prior to the start date of the RT course were considered.

### Clinical Data

The patients’ clinical data included: overall survival, ORNJ status (binary, yes or 1 vs. no or 0), time to event, ORNJ grade, pre-RT dental extractions, T stage, N stage, chemotherapy (induction vs. induction and concurrent vs. concurrent vs. no chemotherapy), post-operative RT vs. definitive RT, HPV/P16 +Ve status (yes vs. no or unknown), tumor site group (oropharynx vs. oral cavity vs. nasopharynx/nasal cavity/paranasal sinuses vs. larynx/hypopharynx vs. major salivary glands vs. other), and mandible volume (in cubic centimeters, cc). 916 patients (81%) were coded with an HPV/P16 +Ve Status of ‘Unknown;’ this missingness of data should be considered in future analyses. Patients with missing data were not included in the original analysis^15^. Overall survival time, binarily coded as 0 for no survival and 1 for survival, represents the time in months between time of RT start date and time of death or time to last follow-up. ORNJ grade was specified using a numeric value of 0-4 following the Tsai staging system^16^; however, in this dataset, ORNJ status was also binarily coded—0 indicated no ORNJ detected and 1 indicated an active ORNJ diagnosis (of any grade) at time of last follow-up. For patients with active ORNJ, time to event is calculated in months from the RT start date to time of ORNJ diagnosis. For patients without an active ORNJ diagnosis, the time to event was censored to be the time in months from RT start date to either time of death or last follow-up. Pre-RT dental extractions were binarily coded—0 indicated negative and 1 indicated positive for pre-RT dental extractions. T stage and N stage indicate the cancer stage, following the standard TNM staging system by the American Joint Committee on Cancer (AJCC, 7^th^/8^th^ edition) and the International Union Against Cancer. Chemotherapy and post-operative RT vs. definitive RT indicate if RT was combined with another treatment; ‘concurrent chemotherapy’ indicates chemotherapy occurred simultaneously with RT while ‘induction chemotherapy’ indicates chemotherapy was completed before RT. Likewise, ‘post-op RT’ indicates RT was completed following surgery while ‘definitive’ indicates RT was completed without surgery. HPV/P16 +Ve indicates positive expression of HPV/P16 via ‘yes’, ‘no’, or ‘unknown.’ Mandible volume was reported (in cc) from delineated mandible contours; mandible bone was auto-segmented with a previously validated multiatlas-based auto-segmentation using commercial software ADMIRE (research version 1.1; Elekta AB, Stockholm, Sweden).

### Dosimetric Data

The patients’ dosimetric data included the following dose-volume metrics: volume of the mandible receiving at least a specified dose (V5-V80 Gy in 5 Gy increments), and dose received by a specified volume of mandible (D0.5%, D1%, D2%, D3%, D5-D95% in 5% increments, D97%, D98%, D99%, D99.5%). These metrics were calculated directly from the radiation dose distribution DICOM files utilizing a Python-based software developed and tested in-house.

#### Data Record

The complete comma-separated value (CSV) file containing demographic, clinical, and dosimetric data for the aforementioned patient population is publicly available at https://doi.org/10.6084/m9.figshare.26240435.v1. This CSV file provides the unique opportunity for analysis of a large HNC cohort with detailed treatment-related information related to prevalence and timing of ORNJ.

#### Technical Validation

Patient demographic and clinical data was stored and accessed via manual extraction by post-doctoral fellows with radiation oncology training from the University of Texas MD Anderson Cancer Center’s Epic Electronic Health Record System server and imported into REDCap electronic data capture tools hosted at the University of Texas MD Anderson Cancer Center^17,18^. Manually pulled data was validated by multiple post-doctoral fellows to minimize misclassification and confirmation biases^19^.

Dosimetric data was obtained from clinical radiotherapy treatment plans using the RayStation treatment planning system (RaySearch Laboratories AB, Stockholm, Sweden). These data were first exported in standardized DICOM-RT (Digital Imaging and Communications in Medicine – Radiation Therapy) format and then analyzed to calculate dose-volume metrics to be used in the model.

**Figure 1.**
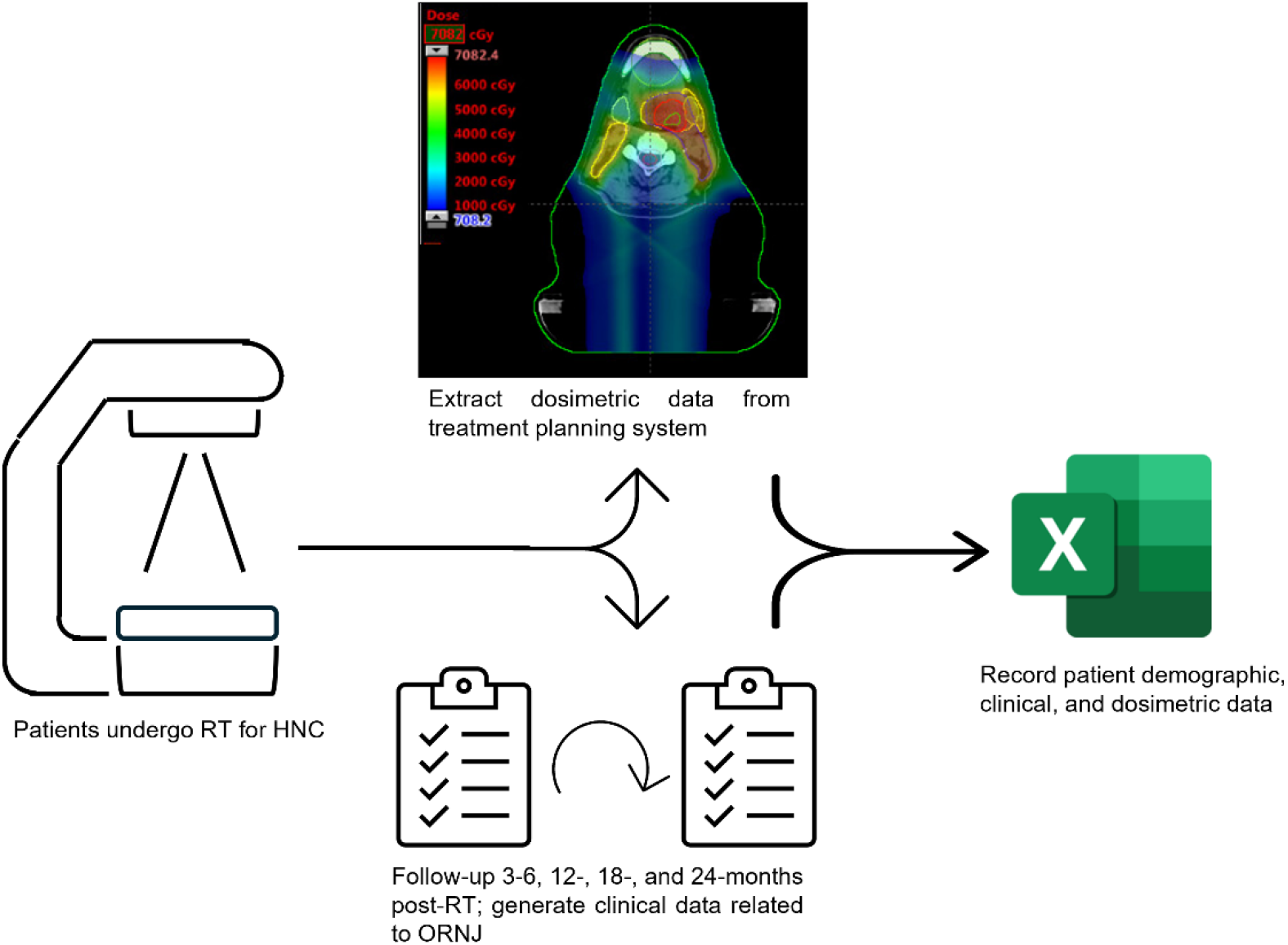
Diagram showing the data collection workflow and input into the final CSV file. Patients underwent RT at MD Anderson Cancer Center (left), in which dosimetric data was generated and acquired from a treatment planning system (top middle). Patient demographic and clinical data were also acquired from initial and follow-up visits (bottom middle). These data were then inputted into the CSV file included within this data descriptor (right).

1471 patients were examined for eligibility for this analysis. 342 patients were excluded due to clinical reasons such as prior irradiations; others were excluded for incomplete or missing data. The final cohort included a dataset of 1129 HNC from MD Anderson Cancer Center.

#### Usage Notes

The CSV file containing all relevant clinical and dosimetric (dose-volume metrics) information is available online on Figshare (https://doi.org/10.6084/m9.figshare.26240435.v1) for further analysis and innovation. Patient identification is anonymized through a randomly assigned subject ID independent from their medical record number (MRN). The WAFT-based time-to-ORNJ online calculator graphical user interface (GUI) is available at https://uic-evl.github.io/OsteoradionecrosisVis/.

## Supporting information

RECORD Checklist

## Data Availability

In accordance with NOT-OD-21-013, Final NIH Policy for Data Management and Sharing, anonymized/de-identified data that support the findings of this study are openly available in an NIH-supported generalist scientific data repository (figshare) at https://doi.org/10.6084/m9.figshare.26240435.v1 no later than the time of an associated publication.
The script used for analyzing this dataset and training and testing the WAFT model can be found here in this repository: https://github.com/LaiaHV-MDACC/ORN-time-to-event-prediction-modelling.

https://github.com/LaiaHV-MDACC/ORN-time-to-event-prediction-modelling

## Code Availability

The script used for analyzing this dataset and training and testing the WAFT model can be found here in this repository: https://github.com/LaiaHV-MDACC/ORN-time-to-event-prediction-modelling.

## Ethics Declarations

After institutional review board approval (RCR030800), data from a philanthropically funded observational cohort at The University of Texas MD Anderson Cancer Center (Stiefel Oropharynx Cancer Cohort, PA14-0947) were extracted for retrospective analysis.

## Consent for Publication

Consent for publication of raw data not obtained but dataset is fully anonymous in a manner that can easily be verified by any user of the dataset. Publication of the dataset clearly and obviously presents minimal risk to confidentiality of study participants (per *BMJ* 2010;340:c181).

## Acknowledgements

NAW is supported by a training fellowship from UTHealth Houston Center for Clinical and Translational Sciences T32 Program (Grant No. T32 TR004905), a NIH National Institute of Dental and Craniofacial Research (NIDCR) Academic Industrial Partnership Grant (R01DE028290), and the American Legion Auxiliary Fellowship in Cancer Research. ZK is supported by a doctoral fellowship from the Cancer Prevention Research Institute of Texas grant RP210042. GEM acknowledges funding support from NIH UG3 TR004501, NSF CNS-2320261, the University Scholar Award from the University of Illinois System, and NIH/NCI R01CA258827. JR received salary support from the NIDCR Diversity Supplement Grant R01DE028290-02S1. KAW was supported by the Image-Guided Cancer Therapy T32 Training Program Fellowship from T32CA261856. KKB acknowledges support from the Image Guided Cancer Therapy Research Program at The University of Texas MD Anderson Cancer Center, which was partially funded by the National Institutes of Health/NCI under award number P30CA016672. MAN received funding from the National Institutes of Health/National Institute of Dental and Craniofacial Research (NIH/NIDCR) through grant R03DE033550. ASRM received funding from NIDCR (U01DE032168, 1R01DE028290-01A1) and NCI (R01CA258827). LVVD received funding and salary support from KWF Dutch Cancer Society through a Young Investigator Grant (KWF-13529) and from NWO ZonMw through the VENI grant (NWO-09150162010173). ACM receives funding from the NIH/NIDCR via grants K12CA088084, R21DE031082, and K01DE030524. CDF, SYL, and KAH receive related funding support from the NIH/NIDCR (U01DE032168). CDF and SYL also receive funding support from the NIH/NIDCR (R01DE025248). CDF also receives infrastructure and salary support through the NIH/NCI MD Anderson Cancer Center Core Support Grant (CCSG) Image-Driven Biologically-informed Therapy (IDBT) program (P30CA016672-47). SYL is supported through the CCSG Head and Neck Program (P30CA016672-48). This work was supported directly or in part by effort, funding, resources or infrastructure from the NIH NCI *OPC SURVIVOR: Optimizing OroPharyngeal Cancer SURVIVORship* Program Project Grant (NCI P01CA285249); and the Charles and Daneen MD Anderson Oropharyngeal Cancer Fund.

## Consortia

### MD Anderson Head and Neck Cancer Symptom Working Group

Natalie A. West, Serageldin Kamel, Zaphanlene Kaffey, Moamen Abdelaal, Melissa M. Chen, Kareem A. Wahid, Jillian Rigert, Kristy K. Brock, Mark Chambers, Adegbenga O. Otun, Ruth Aponte-Wesson, Renjie He, Mohamed A. Naser, Katherine A. Hutcheson, Abdallah S. R. Mohamed, Lisanne V. van Dijk, Amy C. Moreno, Stephen Y. Lai, Clifton D. Fuller, Laia Humbert-Vidan

### Spatial-Non-spatial Multi-Dimensional Analysis of Radiotherapy Treatment/Toxicity Team (SMART3)

Andrew Wentzel, G. Elisabeta Marai, Guadalupe Canahuate, Xinhua Zhang, Abdallah S. R. Mohamed, Clifton D. Fuller

### OPC-SURVIVOR Program

Renjie He, Mohamed A. Naser, Katherine A. Hutcheson, Abdallah S. R. Mohamed, Amy C. Moreno, Stephen Y. Lai, Clifton D. Fuller, Laia Humbert-Vidan

## Author Contributions

Conceptualization: NAW, SK, ZK, MA, GC, XZ, KKB, RH, MAN, KAH, LVVD, ACM, CDF, LHV. Methodology: NAW, SK, ZK, MA, MMC, RH, MAN, KAH, ACM, CDF, LHV. Software: SK, AW, ZK, GEM, CDF, LHV. Validation: SK, CDF, LHV. Formal analysis: SK, ZK, ACM, CDF, LHV. Investigation: NAW, SK, AW, ZK, MA, GEM, GC, XZ, MMC, KAW, JR, KKB, MC, AOO, RAW, RH, MAN, KAH, ASRM, LVVD, ACM, SYL, CDF, LHV. Resources: NAW, SK, ZK, GEM, MAN, KAH, SYL, CDF, LHV. Data curation: NAW, SK, ZK, MA, MMC, CDF, LHV. Writing – original draft: NAW, CDF, LHV. Writing - review and editing: NAW, SK, AW, ZK, MA, GEM, GC, XZ, MMC, KAW, JR, KKB, MC, AOO, RAW, RH, MAN, KAH, ASRM, LVVD, ACM, SYL, CDF, LHV. Visualization: NAW, LHV. Supervision: NAW, SK, ZK, ACM, CDF, LHV. Project administration: NAW, SK, ZK, ACM, CDF, LHV. Funding: KAH, ACM, SYL, CDF.

## Data Citation

https://doi.org/10.6084/m9.figshare.26240435.v1

